# Large Language Model-Driven Prioritization of Alzheimer’s Disease Drug Targets Across Multidimensional Criteria

**DOI:** 10.64898/2025.12.28.25343106

**Authors:** Stanislaw Adaszewski, Torsten Schindler

## Abstract

Large language models (LLMs) offer new opportunities to synthesize the vast and heterogeneous biomedical literature, yet their potential to support drug target prioritization in complex diseases such as Alzheimer’s disease (AD) remains largely unexplored. Here, we introduce an LLM-driven framework that evaluates and ranks AD therapeutic targets across six criteria central to pharmaceutical decision-making: biological confidence, technical feasibility, clinical developability, patient impact, competitive landscape, and safety assessment. Using Gemini 2.5 Pro augmented with real-time web search, we performed large-scale pairwise comparative evaluations and pointwise scoring across a focused set of 522 AD-associated targets with high-quality chemical probes—a tractable subset enriched for clinically advanced targets. We implemented a novel pairwise QuickSort-based ranking procedure that leverages the LLM as a comparative oracle, and benchmarked its performance against pointwise scoring across 16 replicate runs per criterion.

Retrieval-augmented LLM reasoning substantially improved early enrichment of clinically validated AD targets, outperforming LLM-only prompting and approaching the performance of the OpenTargets association benchmark. Pairwise comparative reasoning consistently exceeded pointwise scoring across five of six criteria, yielding higher stability, stronger inter-criterion structure, and markedly improved normalized gain metrics. Multi-objective integration using Pareto fronts and utopia-point scoring further enhanced consensus and robustness, producing holistic rankings that nearly matched the strongest individual criteria while exhibiting superior cross-category coherence. Challenges remained in assessing competitiveness and safety—domains with sparse or inconsistent literature representation—highlighting areas where hybrid models integrating structured datasets may be required.

Together, these results demonstrate that retrieval-augmented LLMs, when combined with structured comparative prompting and multi-criteria integration, can approximate expert-level reasoning and meaningfully enrich target prioritization pipelines for AD. This framework provides a scalable, interpretable, and biologically grounded approach for early-stage drug discovery, with broad applicability to other complex diseases.

**Graphical Abstract:** 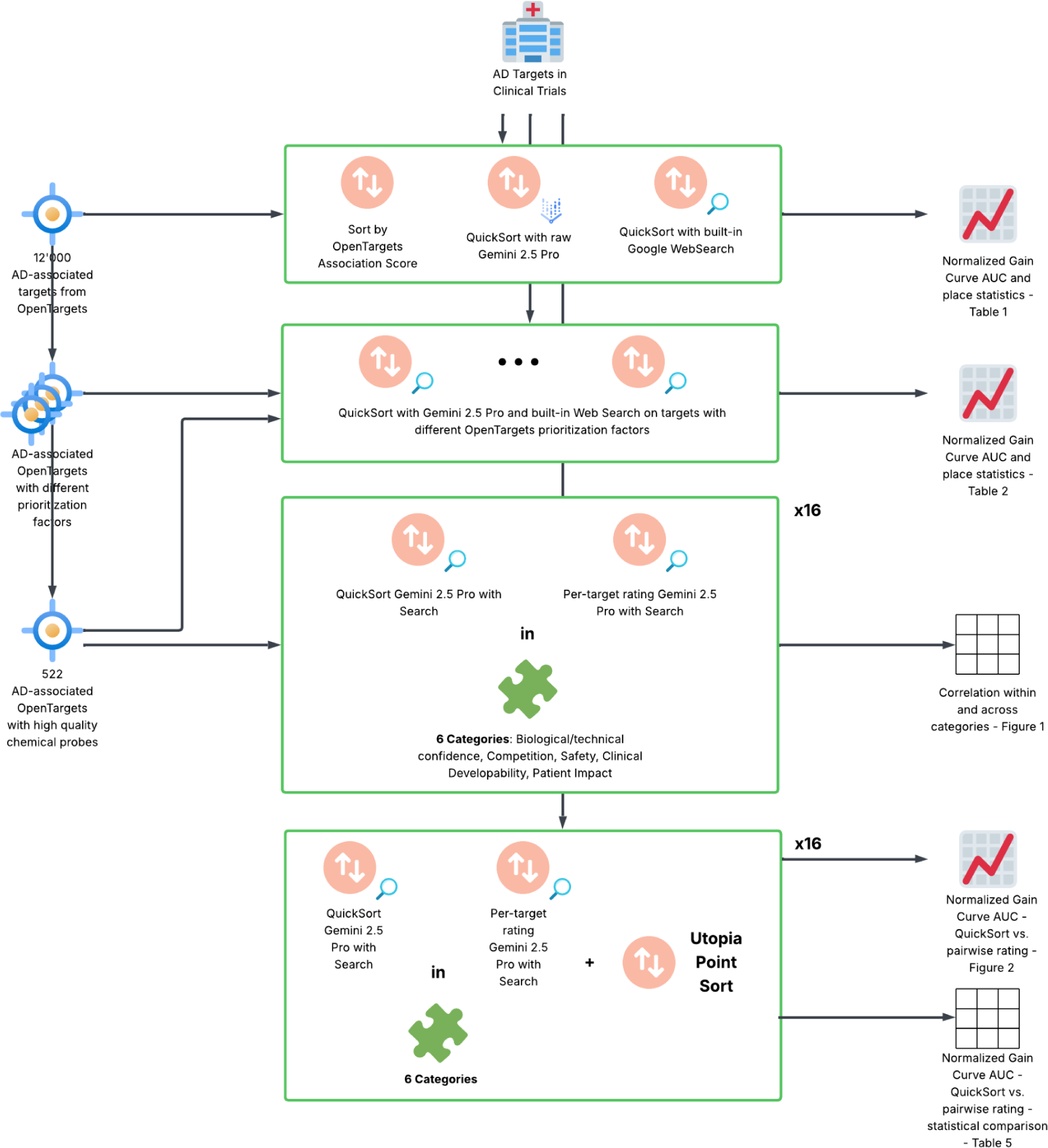

## Introduction

Alzheimer’s disease (AD) – the most common form of dementia – is an urgent global health challenge, rapidly becoming one of the most burdensome and lethal diseases of our time ^1^. Yet despite decades of research, therapeutic progress in AD has been disappointingly slow. Only around 10% of drug candidates that enter clinical development across all diseases ultimately achieve regulatory approval ^2^, and AD drug development in particular has been marked by exceptionally high failure rates (an estimated 99% attrition in the 2002–2012 period) ^3^. To date, available AD treatments provide mostly symptomatic relief, and numerous disease-modifying trials targeting presumed pathological mechanisms have failed due to lack of efficacy or safety issues ^4^. This poor track record reflects the complex, multifactorial etiology of AD – involving amyloid, tau, inflammation, and other processes – and underscores the critical importance of identifying the *right* therapeutic targets amid the myriad of possibilities ^4^. An incorrect or inadequately validated target can doom a drug program from the outset, whereas choosing a biologically compelling target significantly tilts the odds of success in favor of a new therapy (for example, drug mechanisms supported by human genetic evidence are substantially more likely to lead to approved drugs) ^2^.

In the face of these challenges, drug discovery teams have increasingly turned to systematic target prioritization frameworks that integrate diverse lines of evidence to guide decision-making. Recent initiatives have emphasized assembling large-scale data – from genome-wide association studies and functional genomics to omics databases and literature mining – into unified platforms for evaluating target–disease associations ^5^. **Multi-criteria assessment** has emerged as a key principle: rather than relying on any single piece of evidence, researchers score candidate targets across multiple dimensions such as biological plausibility (e.g. genetic or pathophysiological links to AD), tractability or druggability of the target, existence of clinical precedents, potential patient impact, and safety considerations ^6^. By quantifying attributes like these, computational frameworks and expert panels aim to rank targets in a rational, transparent manner, focusing resources on those most likely to yield effective and safe therapies ^5^. Indeed, accumulating evidence suggests that applying such evidence-weighted prioritization can improve the success rate of drug discovery, as seen in the strong correlation between genetically supported targets and later-stage clinical success ^2^. However, current computational approaches for target ranking still face important limitations. Many algorithms (for example, network-based analyses or machine-learning models trained on omics data) capture only subsets of biomedical knowledge and may overlook contextual insights that disease experts glean from the literature ^7^. In addition, the predictive power of these methods is fundamentally constrained by the scope and quality of available training data; for complex diseases like AD, an incomplete understanding of disease biology and sparse “gold-standard” examples of successful targets can hamper the performance of purely data-driven models ^8^. As a result, there remains a significant gap between the rich, qualitative knowledge dispersed in the scientific literature and the quantitative outputs of conventional target prediction pipelines.

**Large language models (LLMs)** have recently emerged as a promising tool to bridge this gap in biomedical knowledge synthesis. LLMs are advanced AI systems trained on massive text corpora, enabling them to comprehend natural language and generate informed responses. In the biomedical domain, LLMs can *read* and *reason* over vast collections of publications, clinical trial descriptions, and database entries, potentially uncovering connections and insights that individual researchers or siloed algorithms might miss. Early studies have demonstrated that general-purpose LLMs like GPT-3 can assist scientists by rapidly analyzing bodies of literature, distinguishing relevant findings across subfields, and even evaluating whether published evidence supports or contradicts a given hypothesis ^9^. This ability to integrate information across disciplines and data sources suggests that LLMs could become powerful allies in complex tasks such as drug target prioritization. In particular, an LLM-based approach could qualitatively evaluate each candidate target through the lens of accumulated biomedical knowledge – for instance, by considering known disease linkages, precedent therapies, molecular tractability, and safety signals reported in the literature – and then provide a reasoned ranking or assessment that mirrors expert judgment. Such an approach offers a fundamentally different yet complementary strategy to traditional algorithms: rather than requiring structured databases or pre-defined training sets for each criterion, an LLM can leverage the *latent knowledge* embedded in unstructured text from millions of publications.

In this study, we explore the use of a state-of-the-art LLM (Gemini 2.5 Pro) to prioritize pharmaceutical targets in AD across six key evaluation categories: **(1) Biological Confidence**, referring to the strength of evidence implicating the target in AD pathogenesis; **(2) Technical Feasibility**, encompassing the tractability of the target (e.g. druggability and ability to modulate it with current technologies); **(3) Clinical Developability**, which considers factors like translational progress or precedents (such as whether molecules against this target have entered trials or shown proof-of-concept); **(4) Patient Impact**, reflecting the potential for meaningful therapeutic benefit on patient outcomes; **(5) Competitive Landscape**, accounting for the novelty of the target and the level of industry/academic activity around it; and **(6) Safety Assessment**, evaluating known or plausible on-target safety risks. These categories collectively span the major dimensions used by pharmaceutical decision-makers when triaging targets, from biological rationale and mechanistic risk, through practical development considerations, to the strategic context of each target. Our approach harnesses the LLM to perform a literature-informed analysis for each criterion: for a given target, the model is prompted with relevant context and asked to provide a judgment (and rationale) for that target’s score or rank in each category. By doing so for a panel of candidate AD targets, we obtain a multidimensional profile for each target that can be aggregated into an overall prioritization. We hypothesize that the LLM’s capacity to synthesize dispersed knowledge will enable a more holistic and up-to-date assessment of AD targets than traditional pipelines, which often rely on static datasets or single-domain evidence.

A central challenge in deploying such an AI-driven target ranking is **validation** of the results. Unlike many tasks in machine learning, here there is no straightforward ground truth – the “ideal” AD drug targets are not definitively known, and outcomes of current hypotheses may remain uncertain for years. This *lack of gold-standard data* complicates the benchmarking of new prioritization methods ^8^. To address this, we propose a comprehensive evaluation framework that triangulates the LLM’s rankings with independent lines of evidence and expert review. Specifically, we assess how stable the set of the LLM’s high-ranked targets is. While such evaluations have inherent limitations in the absence of a true oracle, they provide critical feedback on the behavior of the LLM system. By implementing this framework, we aim to rigorously test the hypothesis that an LLM can perform robust target prioritization and to identify any systematic biases or gaps in its knowledge. In summary, our work introduces a novel LLM-guided approach to AD target ranking, demonstrates its application across multiple prioritization criteria, and lays out a path for its validation in an area where traditional benchmarks are elusive. We envision that this approach, alongside continued refinement and benchmarking, can help accelerate the discovery of high-confidence therapeutic targets for Alzheimer’s disease by leveraging the unparalleled breadth of knowledge encapsulated in modern AI models.

## Methods

### Data and Target Selection

We obtained a comprehensive list of 12,000+ candidate targets associated with Alzheimer’s disease (AD) from the OpenTargets^10^ platform (all targets linked to AD by genetics or literature). As a reference “ground truth” for model evaluation, we compiled a list of known AD drug targets currently in clinical trials (from the supplementary data of ^11^). To focus our experiments, we leveraged OpenTargets’ target annotation factors (also known as “prioritization factors”) – attributes indicating tractability or target readiness (e.g. availability of chemical probes, secreted protein status, presence of ligandable pockets, etc.). Such tractability assessments have been widely used in drug discovery to gauge target “drugability” and development risk ^12^. We analyzed each factor’s coverage and enrichment of known trial targets: for each factor, we computed how many AD-associated targets possess the factor and how many of the known trial targets fall in that subset, and we examined the distribution of trial targets among the top-ranked genes by OpenTargets association score for that factor. This analysis revealed that the *“hasHighQualityChemicalProbe”* factor identified a subset of AD targets that was both sizable and highly enriched in known trial targets (covering 44 known trial targets, more than any other single factor). High-quality chemical probes are recognized as essential tools in target validation and often signal well-studied, tractable targets ^13^. In our data, targets with available high-quality chemical probes showed the greatest overlap with clinical-trial targets, suggesting this feature is a strong indicator of translational progress. By comparison, other factors were less informative (for instance, only 10 known trial targets were secreted proteins and 21 had known druggable pockets). Based on these findings, we restricted subsequent experiments to the 522 AD-associated targets annotated as having high-quality chemical probes. Focusing on this subset strikes a balance between maintaining biological diversity and ensuring a high prior probability of identifying relevant targets, while also keeping the experiment’s scale computationally manageable. All six target-ranking criteria (biological confidence, technical confidence, clinical developability, patient impact, competitiveness, and safety assessment) were considered for these 522 targets (the formal definitions of each criterion are provided in the Introduction, and their prompt formulations are given in the Supplement).

### LLM Model Configuration and Knowledge Augmentation

We employed the **Gemini 2.5 Pro** large language model (an advanced generative text model comparable to GPT-series models) as our ranking engine. All LLM inferences were run in a batch, automated mode to handle the large number of queries required. We evaluated the model in two configurations: *(i)* **Without external knowledge tools**, relying solely on the LLM’s internal knowledge, and *(ii)* **With an integrated web search tool**, which allowed the LLM to perform Google Search queries and retrieve online information during its reasoning. The latter configuration is analogous to retrieval-augmented generation approaches in NLP ^14^, wherein the model can augment its internal knowledge with up-to-date external information. In the context of scientific and factual queries, giving the LLM access to a search tool can significantly improve its accuracy and depth of reasoning ^15^. We first conducted a preliminary experiment to compare these two configurations on the full set of ∼12k AD targets. In this pilot, the model ranked all targets in each category using our LLM-driven QuickSort method (described below), and we computed the ranking’s ability to early-retrieve known trial targets (using the gain curve metric defined later). The web-enabled model consistently achieved higher normalized gain scores than the vanilla model, indicating that on-the-fly retrieval of factual information (e.g. checking gene-disease evidence, known biology, or competitive landscape) helped the LLM make more informed comparisons. Based on this result, all subsequent ranking experiments were performed with the *search-augmented Gemini 2.5 Pro* configuration.

### LLM-Based Target Ranking Approaches

We implemented two distinct ranking methodologies to have the LLM prioritize targets in each of the six criteria: a **pairwise comparison method** using a QuickSort algorithm and a **pointwise scoring method** using independent ratings. Both methods operate in a zero-shot prompting setting (no model fine-tuning), leveraging the LLM’s ability to evaluate targets based on described evidence. All prompts followed a consistent template (provided in the Supplement), with adjustments for each criterion.

#### Pairwise Comparison Ranking with QuickSort

Our primary ranking approach uses the LLM as a comparative oracle to drive a QuickSort procedure. Pairwise comparison prompting has recently emerged as an effective strategy for LLM-based ranking tasks ^16^. Rather than asking the model to assign absolute scores (which can be inconsistent or difficult for the model to calibrate ^16^), the model is asked a simpler question: “Between target A and target B, which is *better* with respect to criterion X?” By only requiring a relative judgment, we reduce the cognitive load on the model and exploit its strength in comparative reasoning. Recent studies have shown that such pairwise ranking prompts enable large language models to achieve ranking performance on par with or exceeding traditional information retrieval systems ^16^.

We embedded these pairwise comparisons within a QuickSort algorithm to efficiently produce a full ordering of all targets for a given criterion. QuickSort is a recursive divide-and-conquer sorting algorithm that selects a *pivot* item and partitions the list into two subsets (“higher” and “lower” priority) based on comparisons to the pivot. In our implementation, the LLM is queried to compare each remaining target to the pivot target (one pairwise comparison at a time), and the model’s judgment (which target is superior in the given category) is used to split the list. The algorithm then recurses on the subsets until a complete ranking is obtained. This approach dramatically reduces the number of pairwise comparisons needed relative to a naïve all-vs-all tournament: QuickSort has an average complexity of *O*(n log n) comparisons, compared to *O*(n²) for all-pairs ranking. In LLM-based ranking, where each comparison is an expensive inference call, minimizing the number of comparisons is crucial ^16^. Prior art in LLM ranking has explored integrating classical sorting algorithms (like HeapSort and BubbleSort) into pairwise prompting frameworks to optimize efficiency ^16,17^. We chose QuickSort for its efficiency and its amenability to parallelization – multiple comparisons (pivot vs several candidates) can potentially be batched in a single LLM call, further reducing overhead. In each comparison prompt, the LLM was provided with concise descriptions of the two target genes (including any relevant details such as known functions, disease associations, or tractability features drawn from OpenTargets and other sources via the search tool) and asked which target better satisfies the criterion in question (for example, *“Which target has higher* biological confidence *for involvement in Alzheimer’s disease, Target A or Target B?”*). The model’s answer (either “A” or “B”, with justification) determined the ordering. By recursively applying this procedure, the model ultimately produces a rank-ordered list of all 522 targets for that criterion.

We note that using an LLM in this pairwise manner implicitly relies on the model’s ability to aggregate diverse pieces of evidence for each target (genetic links, preclinical findings, etc.) and make a holistic judgment. The integrated search tool proved valuable in this context: for example, when comparing two targets for *clinical developability*, the model could fetch current trial information or known druggability data for each protein before deciding. The QuickSort-based ranking is an **innovative element** of our methodology, enabling a human-level comparative evaluation at scale. It is inspired by similar comparative ranking frameworks in other domains (e.g. *pairwise preference learning* and *Elo rating systems*) but to our knowledge this is one of the first applications of QuickSort with an LLM judge in biomedical target prioritization.

#### Pointwise Scoring and Rating-Based Ranking

For comparison, we also implemented a more conventional pointwise approach, in which the LLM evaluates each target independently. In this **rating-based method**, the model is prompted with a single target’s information (again leveraging the search tool for external knowledge) and asked to *rate or score the target* on the given criterion. For instance, the model might be asked: *“On a scale from 1 (very low) to 5 (very high), how would you rate the* technical confidence *of Target X as an Alzheimer’s disease target?”* or *“Briefly assess Target Y’s safety profile and indicate whether its* safety assessment *risk is Low, Medium, or High.”* The exact format of the rating output was tailored to each category (see Supplement for prompt details), but in all cases these ratings were then used to rank the targets from best to worst for that criterion. This pointwise strategy treats the LLM’s outputs as analogous to a human expert’s scoring of each candidate. While straightforward, pointwise ranking with LLMs can be challenging because the model must calibrate an absolute scale across many items; indeed, prior research in LLM ranking has found that zero-shot pointwise judgments often underperform pairwise approaches, as LLMs struggle to maintain consistent criteria across multiple independent queries ^16^.

Nevertheless, this method provides a baseline for comparison and reflects how one might use an LLM in a simpler setting (e.g. to generate a “score” for each gene that could then be sorted). We took care to randomize the order in which targets were presented to the model for scoring, to avoid any ordering biases in generation. Each target’s context included the same standardized data used in the pairwise method (target descriptions, any retrieved evidence, etc.), ensuring the model had comparable information as in the pairwise comparisons.

### Experimental Procedure and Evaluation Metrics

All ranking experiments were conducted on the focused set of 522 AD targets with high-quality chemical probes. We generated rankings for each of the six criteria (biological confidence, technical confidence, clinical developability, patient impact, competitiveness, safety) using both the pairwise QuickSort method and the pointwise rating method described above. To account for variability in LLM responses (due to stochastic sampling in generation or differences in pivot choices in QuickSort), we performed **16 independent runs** of each method for each criterion. Each run of QuickSort was initialized with a different random pivot selection sequence (ensuring the comparison order varied), and each run of the pointwise method re-queried the model fresh for every target, using a different random seed for generation. This yields 16 replicate rankings per (method, criterion) combination, which allowed us to assess the consistency of the rankings and aggregate performance statistics with confidence intervals.

**Performance Metric – Normalized Area Under the Gain Curve:** To evaluate the quality of a given target ranking, we measured how effectively it “ranks high” those targets that are known to be in AD clinical trials (our proxy for likely true positives). We devised a cumulative gain curve analysis for each ranking: as we move down the ranked list from rank 1 to rank *N*, we plot the fraction of known trial targets that have been encountered up to that position (Y-axis) versus the rank position (X-axis). An ideal ranking (one that places all known validated targets at the top) would show a steep curve that quickly reaches 100% of known targets early. A random ranking would, on average, produce a diagonal line (e.g. 50% of known targets found by halfway through the list). We compute the **area under this gain curve** (AUGC) as a summary of early enrichment. To enable comparisons across experiments, we normalize this area to a 0–1 scale by referencing the random expectation and the theoretical maximum. Specifically, if *A<sub>actual</sub>* is the area under the model’s curve, *A<sub>random</sub>* the area under the random baseline curve, and *A<sub>perfect</sub>* the area under the ideal curve (which would capture all known targets immediately at the top), we define:

This normalized metric is 1.0 for a perfect ranking (all trial targets ranked before any others), approximately 0.0 for a random ranking, and can theoretically be negative if the model perversely ranks known targets at the bottom of the list (worse than random). In essence, this metric evaluates the *early recall* of true targets: higher values indicate that the ranking method is successful at bringing known important targets to the forefront. Normalized gain is analogous to evaluation measures used in information retrieval (such as normalized discounted cumulative gain) but here simplified to an un-discounted cumulative recall context for easier interpretability. We calculated this metric for each of the 16 runs per method and category. These values were averaged to compare the **mean performance** of the QuickSort vs. rating-based approach on each criterion, and we also computed standard deviations to gauge variability.

Beyond the primary performance metric, we also analyzed the **consistency and correlation of rankings** across criteria. For each pair of criteria, we evaluated whether a target’s high rank in one tended to imply a high rank in the other. This was done by computing Spearman rank correlation coefficients between the final ranking lists (averaged across runs) for all pairs of criteria. A high correlation would suggest overlap or redundancy between criteria in terms of which targets they prioritize, whereas low or negative correlations would indicate that different criteria favor different targets. This analysis helps understand whether the six dimensions are complementary or if some are surrogates for others in practice. We also examined the run-to-run stability of the rankings for each method: e.g., how often the same top 10 targets appear across the 16 QuickSort runs, to ensure our conclusions are not driven by outlier runs or random LLM fluctuations.

### Multi-Criteria Ranking Integration (Pareto Front and Utopia-Point Method)

Finally, we developed an approach to combine the six individual criteria into an overall prioritization of targets. Treating each criterion as an objective axis, this becomes a **multi-objective ranking problem** in which we seek targets that perform well across all dimensions. Multi-criteria decision analysis techniques are well-suited for such problems and are increasingly used in drug discovery to balance efficacy, safety, and other factors ^18^. We adopted a two-step strategy: (1) **Pareto front analysis**, and (2) **“utopia” point scoring**. In the first step, we identified the Pareto-optimal set of targets: a target is Pareto-optimal (non-dominated) if no other target is strictly better in *all* six criteria. This yields a Pareto ranking (front 1, front 2, etc.), which qualitatively tells us which targets achieve the best trade-offs (front 1) versus those that are dominated by others (lower fronts). However, the Pareto front typically contains many targets (since trade-offs exist and we have no external weights to prefer one criterion over another), so to produce a practical overall ranking, we introduced a “utopia point” method. In this approach, we first define an *ideal* performance point (the utopia): hypothetically, a target that is top-ranked in every category simultaneously. Of course, no real target achieves this ideal, but it provides a reference. We then calculate, for each actual target, a distance to this utopian point in the six-dimensional performance space. In practice, we represented each target by a vector of its percentile ranks in each of the six criteria (so that all dimensions are normalized on a 0–100 scale, where 100 is best). We then computed the Euclidean distance of this vector to the ideal (100,100,100,100,100,100). The targets were then sorted by ascending distance, so that the one closest to the ideal (i.e. most balanced high performance across all criteria) is ranked #1 overall. This *utopia-point ranking* is a form of multi-objective scalarization, conceptually similar to techniques like TOPSIS (Technique for Order Preference by Similarity to Ideal Solution) used in decision analysis ^18^. It provides a single prioritized list of targets that best satisfy *all-round excellence* across the six categories.

We evaluated the effectiveness of this combined ranking in multiple ways. First, we computed the same normalized gain area metric to see if the multi-criteria ranking improved the identification of known trial targets compared to the single-criterion rankings. The expectation was that by integrating complementary criteria (each of which captures different facets of a target’s attractiveness), we might surface targets that are consistently good (though perhaps not top in any one category) and that those might correspond better to real-world successful targets. Second, we examined how the multi-criteria (utopia) ranking compares with each individual category ranking – both in terms of performance and in terms of agreement. We calculated the Spearman correlation between the utopia-based ordering and each of the six single-criterion orderings, to identify which criteria were most aligned with the overall consensus and which were outliers. We also measured how much using a multi-criteria approach trades off performance in any given category: for example, does the utopia-ranked list still achieve high biological confidence and safety, or does it sacrifice one for another? This analysis is akin to checking the *projection* of the utopia ranking onto each criterion’s gain curve. Lastly, as an additional summary, we computed a **Pareto efficiency score** for the utopia ranking: the fraction of utopia-top-ranked targets that lie on the Pareto front (ideally, a good combined ranking would predominantly select Pareto-optimal targets, confirming that it picks up the best trade-offs available).

All computations and statistical analyses were conducted in a Python environment, and the full code along with prompt templates is provided in the Supplement. The methodological choices in our pipeline – from using LLM-driven pairwise sorting to multi-objective combination – were informed by prior research and tailored to the unique challenge of evaluating drug targets with incomplete ground truth. Similar multi-factor prioritization frameworks have been applied in related contexts (e.g., ranking therapeutic antibodies for AD by multiple in vitro criteria ^19^), but to our knowledge this work is the first to leverage a large language model to directly reason over such diverse criteria and perform the ranking task. The following sections (Results and Discussion) detail the outcomes of these methods, comparing the QuickSort vs. rating approaches and analyzing the insights gained from the multi-criteria ranking.

## Results

### 1. Large-Scale LLM-Driven Prioritization of AD Targets

We first evaluated the feasibility and performance of an LLM-driven ranking pipeline by applying Gemini 2.5 Pro to >12,000 AD-associated targets using a high-level “portfolio suitability” prompt. Rankings were generated with and without integrated web search. As summarized in *Table 1*, the search-enabled configuration substantially outperformed the LLM alone, achieving a normalized gain area comparable to the OpenTargets association score benchmark. Specifically, embedding Google Search improved early enrichment of clinical trial targets by ∼80% relative to no-tools prompting, yielding area-under-gain-curve (AUGC) values of 0.72 vs. 0.40, respectively. These results demonstrate that retrieval-augmented reasoning markedly improves the LLM’s ability to surface biologically and clinically relevant AD targets at scale.

**Table 1.** Sort results for 12000+ targets for “portfolio suitability” in Alzheimer’s disease - using LLM without tools and with integrated Web Search embedded in a Quick Sort algorithm.

|  |  |  |  |
| --- | --- | --- | --- |
|  | Without tools | With integrated Web | OpenTargets |

|  |  | Search | Association Score |
| --- | --- | --- | --- |
| **AUC** | 0.40 | 0.72 | 0.73 |
| **min_place** | 0 | 1 | 1 |
| **c25_place** | 850 | 248 | 379 |
| **med_place** | 2583 | 829 | 1325 |
| **c75_place** | 6310 | 2585 | 2243 |
| **max_place** | 12251 | 11067 | 9237 |
| **mean_place** | 3713 | 1779 | 1705 |

### 2. Selection of the High-Quality Chemical Probe Subset

To balance computational tractability with experimental informativeness, we next examined OpenTargets’ prioritization factors and quantified their overlap with known AD clinical trial targets (Table 2). The *hasHighQualityChemicalProbes* category exhibited the highest trial-target enrichment ratio (n₂:n₃ = 44:522) and the lowest normalized gain curve when ranked using OpenTargets alone—suggesting considerable room for methodological improvement. Based on this combination of biological relevance, tractability, and experimental value, we restricted downstream analyses to this subset of 522 targets.

**Table 2.**
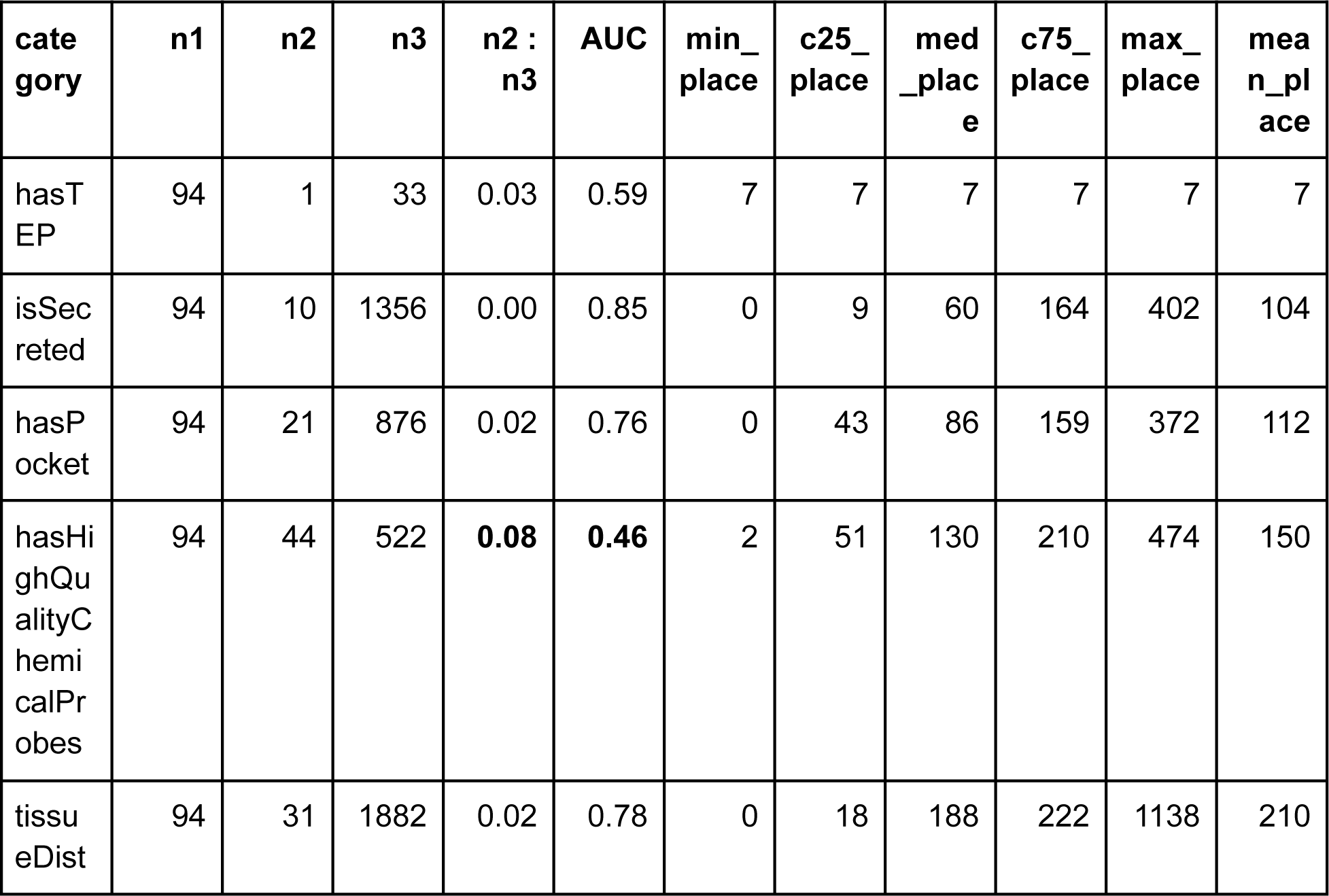
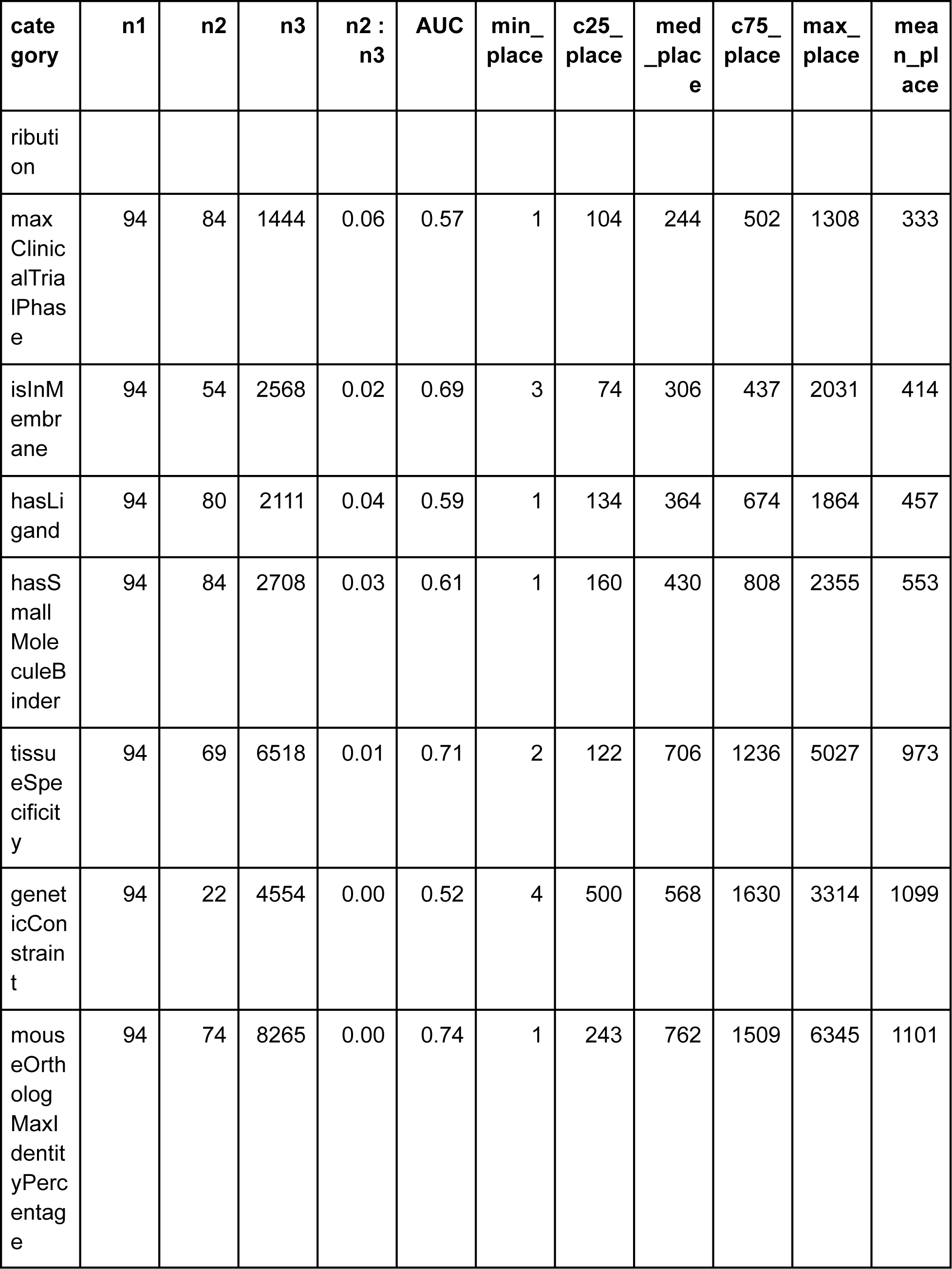
Sorting statistics on 12000+ Alzheimer’s disease targets grouped by OpenTarget prioritization factors. n1, number of targets in clinical trials; n2, number of targets overlapping with the given prioritization factor; n3, number of targets with the given prioritization factor; AUC, normalized area under the gain curve; min_place, highest place; c25_place, 25th centile place; med_place, median place; c75, 75th centile place; max_place, lowest place; mean_place, mean place.

### 3. Pairwise vs. Pointwise Evaluation Across Six Target-Assessment Criteria

We assessed LLM performance under two ranking paradigms—pairwise QuickSort comparisons and pointwise ratings—across six dimensions: biological confidence, technical feasibility, clinical developability, patient impact, competitive landscape, and safety assessment.

#### 3.1. Rank Structure and Inter-Category Correlations

Pairwise rankings exhibited stronger internal consistency and clearer categorical differentiation than pointwise ratings. As shown in *Figure 1a*, biological confidence, clinical developability, and patient impact formed a correlated cluster (Spearman 0.69–0.85), reflecting shared mechanistic and translational evidence bases. Technical confidence and safety assessment displayed moderate correlations with these categories, whereas competitiveness emerged as the least correlated and least stable category across runs.

**Figure 1.**
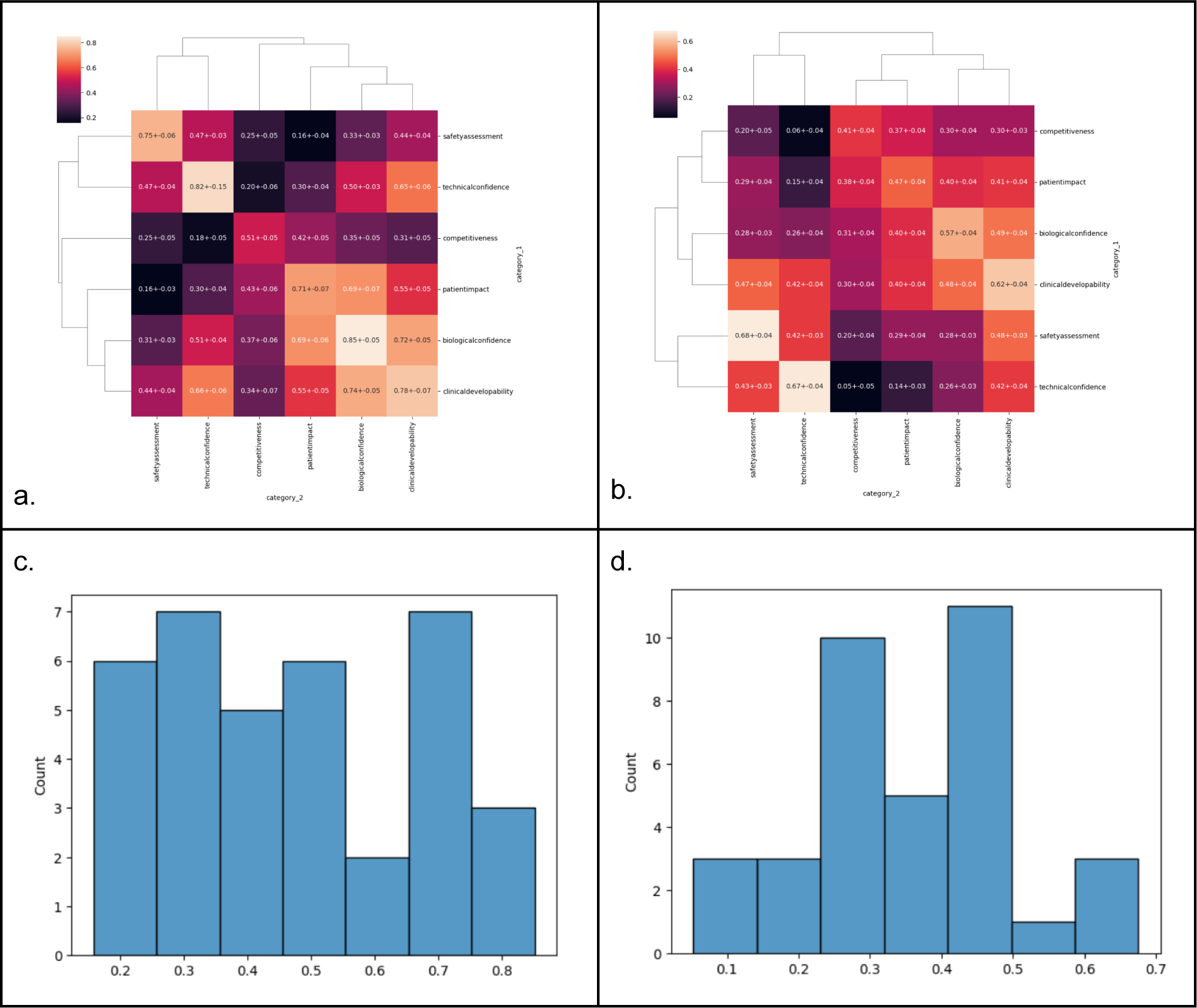
Correlations across categories in pairwise comparison-based rankings (a) and raing-based rankings (b). 40_category_corr_heatmap.ipynb.

In contrast, pointwise scoring yielded reduced within-category stability and attenuated inter-category structure (*Figure 1b*). Correlations tended to homogenize around 0.4–0.5, suggesting less discriminative behavior across dimensions. These shifts imply that pairwise reasoning—and the forced-choice prompt format—encourages more consistent application of criteria than independent rating prompts.

(*Tables 3–4 contain dense correlation matrices and are recommended for placement in the Supplement.*)

#### 3.2. Method Performance Across Criteria

Across five of the six criteria, the pairwise QuickSort method significantly outperformed the pointwise approach in retrieving known trial targets early in the ranking (Figure 2). Large effect sizes were observed for biological confidence (Cohen’s *d* ≈ 6.3), clinical developability (d ≈ 7.1), technical confidence (d ≈ 2.8), and patient impact (d ≈ 2.6). Only competitiveness and safety assessment did not show statistically significant methodological differences (*Table 5*). These findings suggest that the categories grounded in richer mechanistic or translational knowledge benefit most from comparative LLM reasoning, whereas categories that are inherently noisy, ambiguous, or weakly represented in literature (competitiveness, safety) provide less structured signal for LLM evaluation.

**Figure 2.**
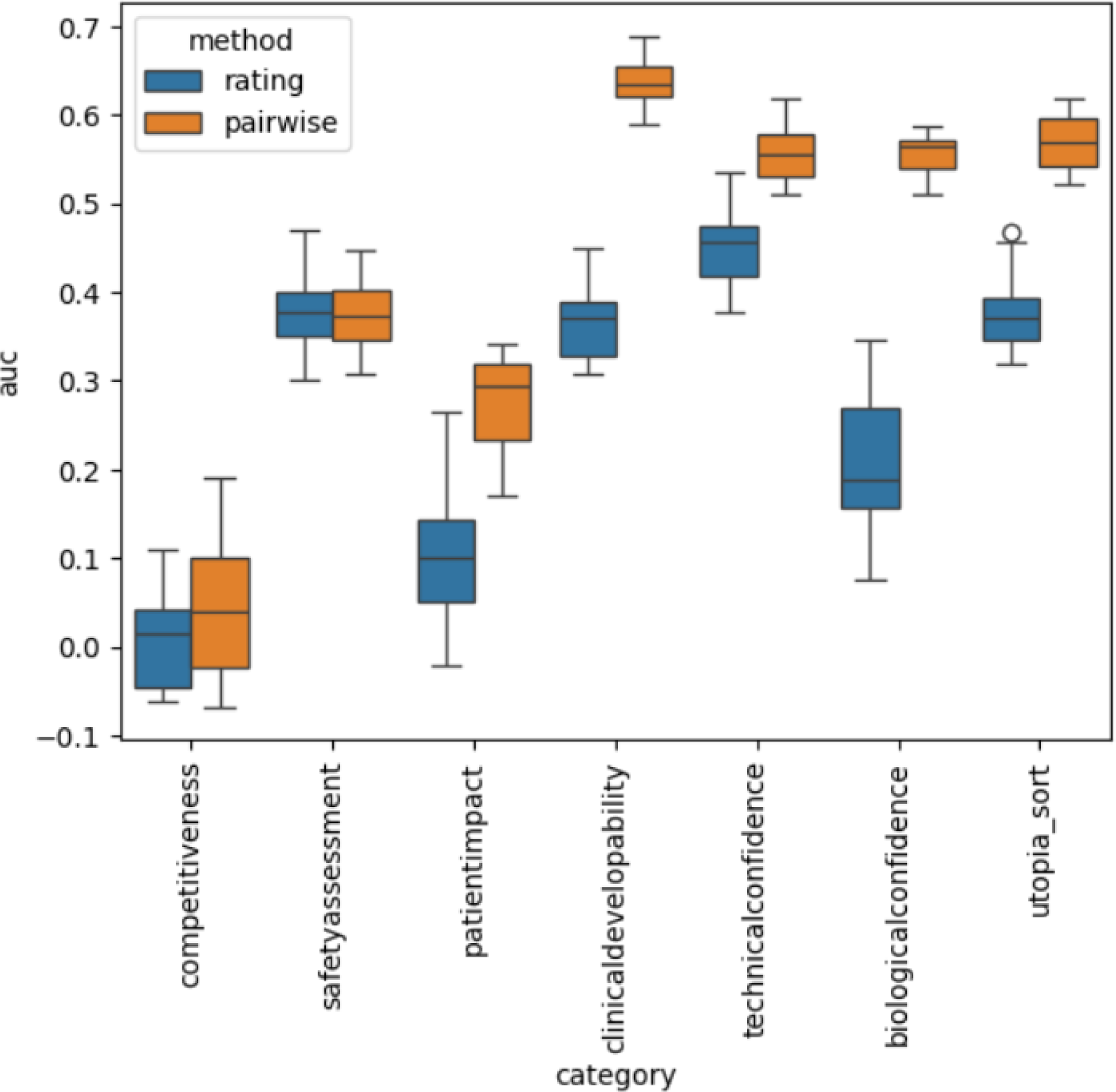
Combined categories and utopia sort comparison. 41_rating_report.ipynb

**Table 5.**
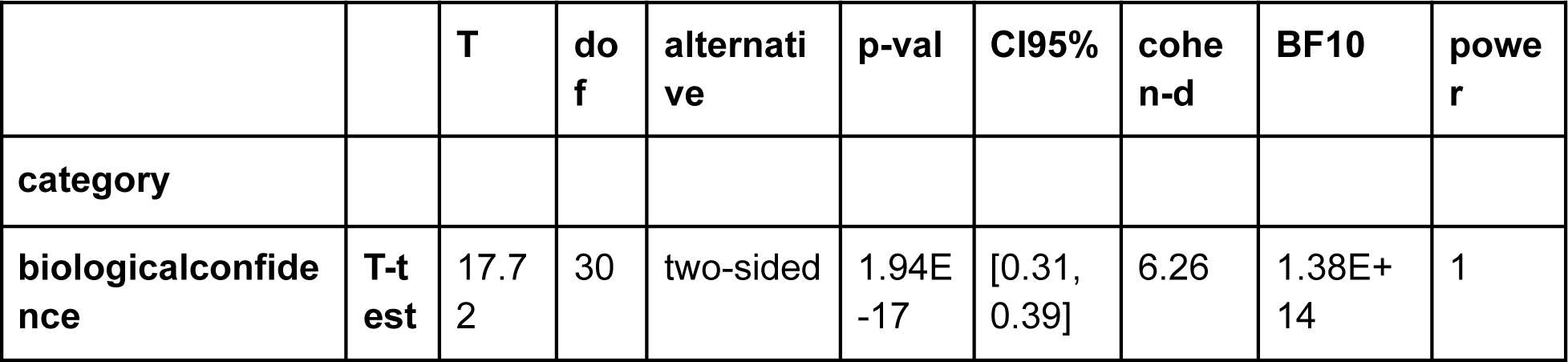
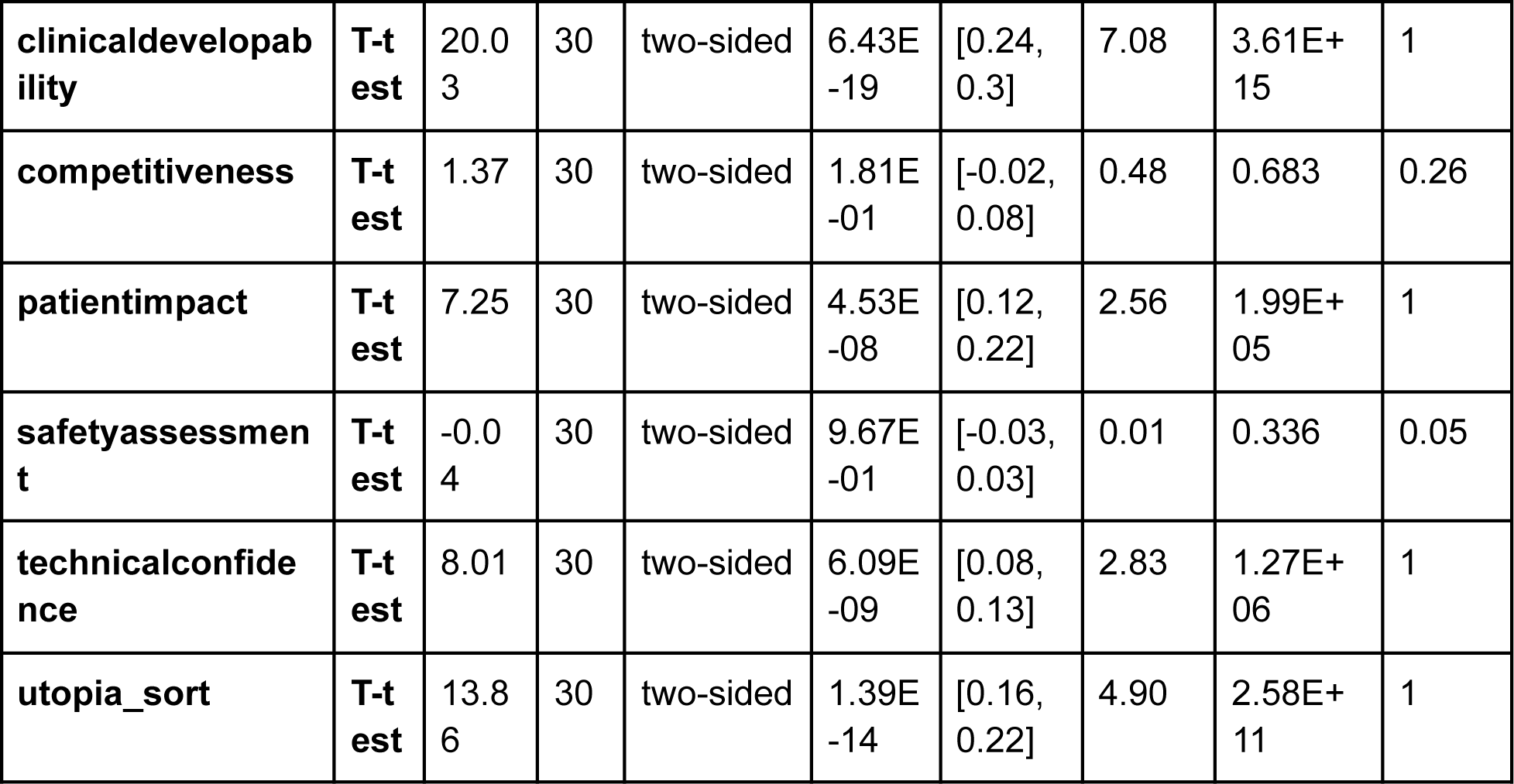
Combined categories and utopia sort comparison. 41_rating_report.ipynb.

|  |  | T | do<br>f | alternati<br>ve | p-val | CI95% | cohe<br>n-d | BF10 | powe<br>r |
| --- | --- | --- | --- | --- | --- | --- | --- | --- | --- |
| category |  |  |  |  |  |  |  |  |  |
| biologicalconfide<br>nce | T-t<br>est | 17.7<br>2 | 30 | two-sided | 1.94E<br>-17 | [0.31,<br>0.39] | 6.26 | 1.38E+<br>14 | 1 |
| **clinicaldevelopability** | **T-test** | 20.03 | 30 | two-sided | 6.43E-19 | [0.24, 0.3] | 7.08 | 3.61E+15 | 1 |
| **competitiveness** | **T-test** | 1.37 | 30 | two-sided | 1.81E-01 | [-0.02, 0.08] | 0.48 | 0.683 | 0.26 |
| **patientimpact** | **T-test** | 7.25 | 30 | two-sided | 4.53E-08 | [0.12, 0.22] | 2.56 | 1.99E+05 | 1 |
| **safetyassessment** | **T-test** | -0.04 | 30 | two-sided | 9.67E-01 | [-0.03, 0.03] | 0.01 | 0.336 | 0.05 |
| **technicalconfidence** | **T-test** | 8.01 | 30 | two-sided | 6.09E-09 | [0.08, 0.13] | 2.83 | 1.27E+06 | 1 |
| **utopia_sort** | **T-test** | 13.86 | 30 | two-sided | 1.39E-14 | [0.16, 0.22] | 4.90 | 2.58E+11 | 1 |

### 4. Multi-Criteria Integration Using Pareto and Utopia-Point Methods

To generate a holistic prioritization, we combined rankings across criteria via Pareto front assignment and utopia-point scoring. The utopia method consistently produced high-performing rankings across runs and categories, achieving normalized gain values nearly matching the best single-criterion models (Figure 2). Moreover, utopia rankings correlated significantly more strongly with individual categories than those categories correlated with one another (*Tables 6–7*), indicating that multi-objective integration enhances stability and consensus.

This pattern was reproduced, though more weakly, in the rating-based approach (*Tables 8–9*). Direct comparison confirmed that utopia-category correlations were significantly higher in the pairwise framework for all criteria except competitiveness and safety (*Table 10*), again reflecting the greater robustness of pairwise reasoning.

Overall, these results demonstrate that multi-criteria integration not only preserves the strengths of individual dimensions but adds orthogonal signal, producing an enriched, consensus-driven ranking suitable for downstream decision-making.

### 5. Summary of Key Findings

- **Retrieval augmentation is essential:** LLM performance with web search approached that of OpenTargets’ association score and substantially exceeded LLM-only performance.
- **Pairwise comparative reasoning outperforms pointwise scoring:** It enhances stability, discriminative ability, and early recall of validated targets.
- **Competitiveness and safety remain challenging categories:** Both exhibit lower stability and weaker separation across methods, likely reflecting sparse or inconsistent evidence in literature.
- **Utopia-point integration yields robust, consensus-aligned rankings:** It performs nearly as well as the strongest individual category but with significantly higher cross-category coherence.
- **The 522-target subset with high-quality probes provides a strong, tractable testbed:** It balances biological relevance with manageable computational cost.

## Discussion

The present study demonstrates that large language models (LLMs), when paired with retrieval augmentation and structured comparative prompting, can perform biologically and translationally meaningful prioritization of Alzheimer’s disease (AD) drug targets. By integrating evidence across six domains central to pharmaceutical decision-making—biological confidence, technical feasibility, clinical developability, patient impact, competitive landscape, and safety assessment—the LLM-based framework captures multidimensional attributes that traditionally require substantial expert time, manual curation, and heterogeneous data integration. The findings highlight both the promise and remaining challenges of deploying LLMs in target prioritization pipelines.

### LLM-driven prioritization complements existing computational approaches

Prior work has shown that target prioritization systems integrating multi-omics datasets and genetic support improve decision-making in drug discovery ^2,5^. However, these systems typically rely on structured databases that cannot fully capture the contextual nuance contained in the scientific literature or clinical trial reports. As elaborated in the Introduction, LLMs offer a fundamentally different way to synthesize dispersed knowledge, overcoming some limitations of network-based methods and data-driven models that lack rich qualitative evidence ^7,8^.

Our results support this hypothesis: retrieval-augmented LLM reasoning achieved normalized gain values approaching those of OpenTargets’ association benchmark (Table 1), far outperforming the same LLM without external search. These findings are consistent with recent demonstrations that retrieval-augmented generation improves factual accuracy and scientific reasoning in expert domains ^14,15^. The ability to cross-reference up-to-date literature, mechanistic studies, and clinical annotations in real time likely enabled the improved identification of known AD trial targets, which often require integration of human genetics, pathological studies, and pharmacological precedent ^1,4^.

### Pairwise comparative reasoning enables more reliable ranking than pointwise scoring

A core result of the study is that pairwise comparative reasoning—implemented through an LLM-driven QuickSort procedure—substantially outperforms pointwise scoring across nearly all criteria (Figure 2). This observation aligns with emerging findings in the LLM literature demonstrating that relative judgments reduce calibration drift and mitigate inconsistencies inherent to independent scoring ^16,17^.

Biological confidence, technical feasibility, clinical developability, and patient impact all showed strong effect sizes favoring pairwise comparison, reflecting that these categories rely on integrated mechanistic and translational evidence, which LLMs are better able to contrast than to quantify independently. The clustering of these categories in Figure 1a suggests that they tap into related evidence streams, including genetics, preclinical findings, and therapeutic precedent—domains where expert reasoning has historically been most predictive of drug development success ^2^.

In contrast, competitiveness and safety assessment remained challenging. Sparse, inconsistent, or poorly structured literature in these areas likely limits what LLMs can robustly infer. Safety in particular is notoriously difficult to evaluate prospectively, as most risk emerges only in late-stage development or real-world evidence ^6^. Competitiveness depends on dynamic, often proprietary pipelines and industry activity that may be incompletely represented even in public-facing sources. Future applications may benefit from supplementing LLM reasoning with structured industry datasets or pharmacovigilance signals (e.g., FAERS, OpenFDA).

### Implications of the high-quality chemical probe subset

Restricting analyses to the 522 AD-associated targets with high-quality chemical probes enabled a tractable yet representative benchmark enriched for biologically and translationally mature targets (Table 2). Chemical probes are strong indicators of experimental readiness and mechanistic tractability ^12,13^; their enrichment among known AD trial targets further validates their utility as a focused discovery set.

From a methodological perspective, focusing on this subset allowed systematic evaluation of ranking methods without incurring prohibitive computational costs. As the Methods outline, full all-pairs comparisons across thousands of targets would be orders of magnitude more expensive. While next-generation LLM systems may reduce inference cost or enable batching of pairwise reasoning, the present findings emphasize the need for hierarchical or staged ranking strategies—e.g., combining coarse-grained filtering with high-resolution pairwise evaluation—for industrial-scale prioritization.

### Multi-criteria integration enhances stability, consensus, and translational relevance

The multi-objective integration using Pareto fronts and utopia-point scoring produced a ranking that nearly matched the best-performing single criteria while substantially improving cross-category coherence (Figure 2 and Tables 6–7). This behaviour reflects classical multi-criteria decision analysis principles, in which consensus solutions often outperform single-objective optima by balancing competing dimensions of developability, tractability, and risk ^18^.

The utopia-point ranking correlates strongly with biological, technical, clinical, and patient impact categories (Tables 6–7), suggesting that these dimensions collectively capture high-value AD target attributes. Importantly, utopia-point integration mitigated noise in categories with lower signal—competitiveness and safety—by anchoring the overall ranking to criteria that consistently displayed strong internal structure (Figure 1a). This finding parallels observations in recent multi-criteria ranking frameworks where uncertain categories benefit from anchoring to more reliable dimensions ^19^ and may reflect a broader principle applicable in other therapeutic areas.

### Limitations and considerations for practical deployment

Despite promising results, several limitations merit discussion.

#### 1. Absence of definitive ground truth

As emphasized in the Introduction, AD target validation lacks a true gold standard. Clinical trial progression is an imperfect proxy, constrained by historical biases, commercial priorities, and incomplete biological understanding ^4,11^. Consequently, gain-curve metrics provide useful—but indirect—assessments of ranking success.

#### 2. Literature bias and LLM hallucination risks

LLMs inherit biases from their training corpora and may over-weight well-studied pathways (e.g., amyloid, tau), mirroring the literature’s historical concentration ^1^. Although retrieval augmentation mitigates hallucination by grounding responses in verifiable sources ^14,15^, the model may still reason confidently from incomplete or skewed evidence. Integrating structured quantitative datasets—genome-wide association results, proteomics, or perturbation screens—may further stabilize reasoning in future applications.

#### 3. Challenges in safety and competitive landscape assessment

Safety prediction requires sophisticated modeling of pathway biology, off-target activity, and toxicological mechanisms—elements not reliably inferable from text alone. Similarly, competitiveness requires near-real-time awareness of the therapeutic pipeline. Both may require hybrid models combining LLM reasoning with dedicated safety/toxicology prediction tools (e.g., structural alerts, mechanistic toxicology models) or curated commercial intelligence databases.

#### 4. Computational scaling

As noted in the draft Discussion and supported by our experiments on the full 12,000+ target set, pairwise approaches scale superlinearly in cost. Emerging setwise-ranking strategies for LLMs ^17^ and approximate tournament methods (e.g., Elo-style adaptive comparison) may reduce inference overhead. Additionally, future work might explore active-learning strategies to select only the most informative comparisons.

### Future directions

Several opportunities emerge from this work.

**Integration with quantitative datasets:** Hybrid models linking mechanistic LLM reasoning with genetic association effect sizes, multimodal omics, or causal network inference may yield improved biological confidence assessments.

**Iterative co-evaluation with experts:** Similar to human-AI collaborative frameworks in other biomedical areas, combining LLM-derived rankings with expert adjudication may enhance reproducibility and reduce risk of systematic biases.

**Application to other diseases:** The framework is generalizable to complex diseases beyond AD, especially those with heterogeneous mechanistic bases (e.g., Parkinson’s disease, ALS), where multi-dimensional prioritization is equally critical.

**Automated portfolio simulation:** Embedding LLM-based rankings into end-to-end target triage workflows—including risk-adjusted portfolio modeling or go/no-go decision support—could bring practical utility to early-stage R&D.

## Conclusions

This study demonstrates that LLMs equipped with retrieval augmentation and structured comparative reasoning can approximate expert-level target prioritization in AD, outperforming pointwise scoring and yielding rankings enriched for clinically validated targets. Multi-criteria integration further enhances stability and consensus, providing a pragmatic, interpretable framework for target triage. While limitations remain—particularly in safety and competitiveness assessment—the evidence indicates that LLM-driven prioritization can meaningfully complement existing computational pipelines and expert workflows. Continued methodological refinement, incorporation of diverse data modalities, and rigorous prospective validation will be essential to realizing the full potential of LLMs in accelerating drug discovery.

## Data Availability

All data produced in the present study are available upon reasonable request to the authors.

## Notes

### Competing Interest Statement

The authors have declared no competing interest.

### Funding Statement

This study did not receive any funding

