## Supplement for "Large Language Model-Driven Prioritization of Alzheimer’s Disease Drug Targets Across Multidimensional Criteria"

Table 3. How correlated the categories are between each other and between utopia (Spearman correlation of positions of a target in rankings based on respective categories)

| y | biologicalconfidence | clinicaldevelopability | competitiveness | patientimpact | safetyassessment | technicalconfidence | utopia |
|---|---|---|---|---|---|---|---|
| x | | | | | | | |
| biologicalconfidence | NaN | 0.967981 | 0.835965 | 0.968950 | 0.777223 | 0.897716 | 0.978383 |
| clinicaldevelopability | 0.967684 | NaN | 0.816273 | 0.934318 | 0.879451 | 0.947033 | 0.983642 |
| competitiveness | 0.815841 | 0.792279 | NaN | 0.873245 | 0.706592 | 0.562327 | 0.924439 |
| patientimpact | 0.962501 | 0.928235 | 0.874736 | NaN | 0.540394 | 0.774019 | 0.962553 |
| safetyassessment | 0.782930 | 0.879755 | 0.716835 | 0.548607 | NaN | 0.891533 | 0.942160 |
| technicalconfidence | 0.879497 | 0.939544 | 0.580587 | 0.750925 | 0.881092 | NaN | 0.954636 |
| utopia | 0.979582 | 0.982870 | 0.929834 | 0.964642 | 0.938481 | 0.963989 | NaN |

Table 4. How correlated performance (AUC under normalized gain curve) is between categories

| category_2 | biologicalconfidence | clinicaldevelopability | competitiveness | patientimpact | safetyassessment | technicalconfidence |
|---|---|---|---|---|---|---|
| **category_1** | | | | | | |
| biologicalconfidence | 0.85+-0.05 | 0.72+-0.05 | 0.37+-0.06 | 0.69+-0.06 | 0.31+-0.03 | 0.51+-0.04 |
| clinicaldevelopability | 0.74+-0.05 | 0.78+-0.07 | 0.34+-0.07 | 0.55+-0.05 | 0.44+-0.04 | 0.66+-0.06 |
| competitiveness | 0.35+-0.05 | 0.31+-0.05 | 0.51+-0.05 | 0.42+-0.05 | 0.25+-0.05 | 0.18+-0.05 |
| patientimpact | 0.69+-0.07 | 0.55+-0.05 | 0.43+-0.06 | 0.71+-0.07 | 0.16+-0.03 | 0.30+-0.04 |
| safetyassessment | 0.33+-0.03 | 0.44+-0.04 | 0.25+-0.05 | 0.16+-0.04 | 0.75+-0.06 | 0.47+-0.03 |
| technicalconfidence | 0.50+-0.03 | 0.65+-0.06 | 0.20+-0.06 | 0.30+-0.04 | 0.47+-0.04 | 0.82+-0.15 |

Table 6. Correlation coefficient of rankings between categories and the utopia approach in the pairwise method. 42_cat_vs_cat_place_both.ipynb

| category_2 | biologicalconfidence | clinicaldevelopability | competitiveness | patientimpact | safetyassessment | technicalconfidence | utopia |
|---|---|---|---|---|---|---|---|
| **category_1** | | | | | | | |
| biologicalconfidence | - | 0.73±0.06 | 0.36±0.06 | 0.69±0.07 | 0.32±0.03 | 0.50±0.04 | 0.80±0.05 |

| | | | | | | | |
|---|---|---|---|---|---|---|---|
| clinicaldevelopability | 0.73±0.06 | - | 0.33±0.06 | 0.55±0.04 | 0.44±0.05 | 0.66±0.06 | 0.82±0.06 |
| competitiveness | 0.36±0.06 | 0.33±0.06 | - | 0.44±0.06 | 0.25±0.05 | 0.19±0.05 | 0.58±0.07 |
| patientimpact | 0.69±0.07 | 0.55±0.04 | 0.44±0.06 | - | 0.16±0.05 | 0.31±0.04 | 0.70±0.06 |
| safetyassessment | 0.32±0.03 | 0.44±0.05 | 0.25±0.05 | 0.16±0.05 | - | 0.48±0.05 | 0.61±0.05 |
| technicalconfidence | 0.50±0.04 | 0.66±0.06 | 0.19±0.05 | 0.31±0.04 | 0.48±0.05 | - | 0.70±0.06 |
| utopia | 0.80±0.05 | 0.82±0.06 | 0.58±0.07 | 0.70±0.06 | 0.61±0.05 | 0.70±0.06 | - |

Table 7. Statistical comparison of correlation between category_1 and category_2 and utopia method and category_2 respectively in the pairwise approach. Down arrow indicates that correlation (category_1, category_2) is lower than (utopia, category_2). Stars indicate Bonferroni-corrected significance level, *** < 0.001, ** < 0.01.
notebooks/42_cat_vs_cat_place.ipynb

| **category_2** | **biologicalconfidence** | **clinicaldevelopability** | **competitiveness** | **patientimpact** | **safetyassessment** | **technicalconfidence** |
|---|---|---|---|---|---|---|
| **category_1** | | | | | | |
| biologicalconfidence | - | ↓ *** | ↓ *** | ↓ | ↓ *** | ↓ *** |
| clinicaldevelopability | ↓ *** | - | ↓ *** | ↓ *** | ↓ *** | ↓ ** |
| competitiveness | ↓ *** | ↓ *** | - | ↓ *** | ↓ *** | ↓ *** |
| patientimpact | ↓ *** | ↓ *** | ↓ *** | - | ↓ *** | ↓ *** |
| safetyassessment | ↓ *** | ↓ *** | ↓ *** | ↓ *** | - | ↓ *** |

| | | | | | | |
|---|---|---|---|---|---|---|
| technicalconfidence | ↓ *** | ↓ *** | ↓ *** | ↓ *** | ↓ *** | - |

Table 8. Correlation coefficient of rankings between categories and the utopia approach in the rating method. 42_cat_vs_cat_place_both.ipynb

| **category_2** | **biologicalconfidence** | **clinicaldevelopability** | **competitiveness** | **patient impact** | **safetyassessment** | **technicalconfidence** | **utopia** |
|---|---|---|---|---|---|---|---|
| **category_1** | | | | | | | |
| biologicalconfidence | - | 0.48±0.04 | 0.31±0.03 | 0.39±0.03 | 0.28±0.03 | 0.27±0.04 | 0.61±0.04 |
| clinicaldevelopability | 0.48±0.04 | - | 0.30±0.04 | 0.40±0.04 | 0.47±0.04 | 0.42±0.03 | 0.69±0.05 |
| competitiveness | 0.31±0.03 | 0.30±0.04 | - | 0.37±0.05 | 0.20±0.04 | 0.05±0.04 | 0.57±0.05 |
| patientimpact | 0.39±0.03 | 0.40±0.04 | 0.37±0.05 | - | 0.29±0.03 | 0.14±0.03 | 0.65±0.04 |
| safetyassessment | 0.28±0.03 | 0.47±0.04 | 0.20±0.04 | 0.29±0.03 | - | 0.42±0.04 | 0.63±0.04 |
| technicalconfidence | 0.27±0.04 | 0.42±0.03 | 0.05±0.04 | 0.14±0.03 | 0.42±0.04 | - | 0.53±0.04 |
| utopia | 0.61±0.04 | 0.69±0.05 | 0.57±0.05 | 0.65±0.04 | 0.63±0.04 | 0.53±0.04 | - |

Table 9. Statistical comparison of correlation between category_1 and category_2 and utopia method and category_2 respectively in the rating approach. Down arrow indicates that correlation (category_1, category_2) is lower than (utopia, category_2). Stars indicate Bonferroni-corrected significance level, *** < 0.001, ** < 0.01. notebooks/42_cat_vs_cat_place.ipynb

| category_2 | biologicalconfidence | clinicaldevelopability | competitiveness | patientimpact | safetyassessment | technicalconfidence |
|---|---|---|---|---|---|---|
| **category_1** | | | | | | |
| biologicalconfidence | - | ↓ *** | ↓ *** | ↓ *** | ↓ *** | ↓ *** |
| clinicaldevelopability | ↓ *** | - | ↓ *** | ↓ *** | ↓ *** | ↓ *** |
| competitiveness | ↓ *** | ↓ *** | - | ↓ *** | ↓ *** | ↓ *** |
| patientimpact | ↓ *** | ↓ *** | ↓ *** | - | ↓ *** | ↓ *** |
| safetyassessment | ↓ *** | ↓ *** | ↓ *** | ↓ *** | - | ↓ *** |
| technicalconfidence | ↓ *** | ↓ *** | ↓ *** | ↓ *** | ↓ *** | - |

Table 10. Comparison of correlation of the utopia ranking with the other categories in respectively pairwise and rating method. The up arrow indicates that the correlation is higher in the pairwise method.

| category_2 | t | pval | df | pval_corr | formatted |
|---|---|---|---|---|---|
| biologicalconfidence | 27.01 | 3.91E-14 | 15 | 2.35E-13 | ↑ *** |
| clinicaldevelopability | 15.54 | 1.18E-10 | 15 | 7.06E-10 | ↑ *** |
| competitiveness | 0.49 | 6.31E-01 | 15 | 3.78E+00 | ↑ |
| patientimpact | 4.40 | 5.14E-04 | 15 | 3.08E-03 | ↑ ** |
| safetyassessment | -1.96 | 6.89E-02 | 15 | 4.13E-01 | ↓ |
| technicalconfidence | 19.65 | 4.06E-12 | 15 | 2.44E-11 | ↑ *** |
